# Mechanisms of Arrhythmogenicity in Hypertrophic Cardiomyopathy: Insight from Noninvasive Electrocardiographic Imaging

**DOI:** 10.1101/19002782

**Authors:** Erick A. Perez-Alday, Kazi T. Haq, David M. German, Christopher Hamilton, Kyle Johnson, Francis Phan, Nichole M. Rogovoy, Katherine Yang, Ashley Wirth, Jason A.Thomas, Khidir Dalouk, Cristina Fuss, Maros Ferencik, Stephen Heitner, Larisa G. Tereshchenko

## Abstract

**Background:** Mechanisms of arrhythmogenicity in hypertrophic cardiomyopathy (HCM) are not well understood.

**Objective:** To characterize an electrophysiological substrate of HCM in comparison to ischemic cardiomyopathy (ICM), or healthy individuals.

**Methods:** We conducted a prospective case-control study. The study enrolled HCM patients at high risk for ventricular tachyarrhythmia (VT) (n=10; age 61±9 y; left ventricular ejection fraction (LVEF) 60±9%), and three comparison groups: healthy individuals (n=10; age 28±6 y; LVEF>70%), ICM patients with LV hypertrophy (LVH) and known VT (n=10; age 64±9 y; LVEF 31±15%), and ICM patients with LVH and no known VT (n=10; age 70±7y; LVEF 46±16%). All participants underwent 12-lead ECG, cardiac CT or MRI, and 128-electrode body surface mapping (BioSemi ActiveTwo, Netherlands). Non-invasive voltage and activation maps were reconstructed using the open-source SCIRun (University of Utah) inverse problem-solving environment.

**Results:** In the epicardial basal anterior segment, HCM patients had the greatest ventricular activation dispersion [16.4±5.5 vs. 13.1±2.7 (ICM with VT) vs. 13.8±4.3 (ICM no VT) vs. 8.1±2.4 ms (Healthy); P=0.0007], the largest unipolar voltage [1094±211 vs. 934±189 (ICM with VT) vs. 898±358 (ICM no VT) vs. 842±90 µV (Healthy); P=0.023], and the greatest voltage dispersion [median(interquartile range) 215(161-281) vs. 189(143-208) (ICM with VT) vs. 158(109-236) (ICM no VT) vs. 110(106-168)µV (Healthy); P=0.041]. Differences were also observed in other endo-and epicardial basal and apical segments.

**Conclusion:** HCM is characterized by a greater activation dispersion in basal segments, a larger voltage, and a larger voltage dispersion through LV.

## Introduction

Patients with hypertrophic cardiomyopathy (HCM) are at high risk of life-threatening ventricular arrhythmias and sudden cardiac death (SCD).^1^ Mechanisms of arrhythmogenicity in HCM are complex and incompletely understood. It was previously shown that the late sodium current is increased in HCM, suggesting the importance of repolarization abnormalities.^2^ At the same time, cardiac magnetic resonance (CMR) studies have shown that the myocardium in HCM is characterized by increased fibrosis burden, supporting an alternative mechanism for arrhythmogenesis - heterogeneity in electrical activation. The degree of late gadolinium enhancement in HCM is associated with SCD and appropriate implantable cardioverter-defibrillator (ICD) therapy.^3^ While the presence of a patchy scar in HCM suggests likely similarity with macro-reentrant post-myocardial infarction (MI) ventricular tachycardia (VT) mechanisms, VT ablation in HCM is less successful than in post-infarction VT. In HCM patients who underwent VT ablation, the incidence of VT recurrence, death, and cardiac transplantation at one year was one of the highest amongst all non-ischemic cardiomyopathies (NICM),^4^ even after adjusting for comorbidities. This may be due to anatomic limitations for ablation (predominantly mid-myocardial septal location of the scar), or diffuse nature of cardiomyocyte disarray and interstitial fibrosis that is the histopathological hallmark of HCM^5^. By and large, the electrophysiological (EP) substrate in HCM is incompletely understood. Recently, the non-invasive electrocardiographic imaging (ECGi), a state-of-the-art technology, became available as a tool to study mechanisms of cardiac arrhythmias.^6^ We designed this study with the goal to describe the EP substrate of HCM, in comparison with the relatively well-understood EP substrate of post-infarction macro-reentrant VT.

## Methods

### Study population: inclusion and exclusion criteria

We conducted a single-center case-control study of high-risk HCM cases with three comparison groups (Clinical Trial Registration—URL: www.clinicaltrials.gov Unique identifier: NCT02806479). The Oregon Health & Science University (OHSU) Institutional Review Board (IRB) approved this study, and all participants signed an informed consent form. Enrollment was performed at OHSU in 2016-2018. Adult (age≥18y) non-pregnant participants were enrolled if the inclusion and exclusion criteria were met, as described below.

Inclusion criteria for HCM group were: (1) a history of resuscitated sudden cardiac arrest, or documented sustained VT, or (2) a maximal left ventricular (LV) wall thickness above 30 mm, or extensive fibrosis on CMR (above 10% of total myocardial volume), or (3) high risk of SCD (>7.5%/5y) as determined by HCM risk-SCD^7^ score.

Healthy control group I (*Healthy*) was designed to include individuals who were free from structural heart disease and arrhythmogenic substrate in ventricles. The inclusion criterion required evaluation by a cardiac electrophysiologist for AV nodal reentrant tachycardia.

Exclusion criteria were diagnosed structural heart disease, or known risk factors of structural heart disease^8^ (history of hypertension, smoking, diabetes, body mass index < 18.5 or >30 kg/m^2^, and family history of coronary heart disease (CHD) diagnosed at age 50 or younger).

Group II (*post-MI VT-free*) included post-MI VT-free patients as documented by at least one generator life of primary prevention ICD, or medical record.

Group III (*post-MI VT*) included post-MI patients with the history of sudden cardiac arrest and implanted secondary prevention ICD, or, if an ICD was implanted for primary prevention of SCD and there was documented sustained (cycle length < 240ms) VT treated by appropriate ICD shock. MADIT-RIT programming criteria were applied, to avoid inclusion of treated non-sustained VT events. Sudden cardiac arrest due to a transient cause was an exclusion criterion.

In addition, exclusion criteria for all study participants were the age of less than 18y, pregnancy, persistent atrial fibrillation (AF), chronic (above 5%) right ventricular (RV) or biventricular pacing, renal insufficiency with estimated glomerular filtration rate (eGFR) < 30 ml/min, congenital heart disease, and contraindications for CMR or cardiac computed tomography (CT) with contrast. By design, we planned to enroll 10 participants in each group.

### Cardiac imaging and assessment of cardiac structure and function

Healthy controls underwent non-contrast CMR using a Siemens TIM Trio 3 Tesla with VB17 software and Siemens Prisma Fit 3 Tesla scanner with E11C software. The other three groups underwent prospectively ECG-triggered contrast-enhanced 256-detector row cardiac CT (Philips iCT, Philips Medical Imaging, Cleveland, OH). The images were acquired in mid to end diastole with a slice thickness of 0.6 mm and in-plane resolution of approximately 0.5 mm. A cardiologist (DMG) reviewed all cardiac CT and MRI images, and ventricular volumes were obtained in a semiautomatic fashion using commercially available software (IntelliSpace Portal; Philips Healthcare, Redmond, WA, USA; and CVI42; Circle Cardiovascular Imaging Inc., Calgary, Alberta, Canada). The standardized myocardial segmentation and nomenclature^9^ were used to define 17 segments of LV. For subjects who underwent CMR, left ventricular ejection fraction (LVEF) was calculated from ventricular volume measurements.

Additionally, data from the most recent echocardiogram and CMR was abstracted to provide additional information on baseline cardiac structure and function. LVEF was calculated from the echocardiogram using the biplane Simpson method of discs. Regional LV function was evaluated by the echocardiographic wall motion score index. Motion and systolic thickening in each segment was scored as: normal or hyperkinesis = 1, hypokinesis = 2, akinesis = 3, and dyskinesis (or aneurysmal) = 4. The wall motion score index was calculated as the sum of all scores divided by the number of visualized segments. The resting peak LVOT gradient was calculated for all participants. In addition, HCM participants had peak LVOT gradient measured during Valsalva maneuver and at peak exertion. The location of fibrosis on CMR images was recorded. Age-and sex-specific thresholds were used to define left ventricular hypertrophy (LVH) on cardiac CT and CMR^10^.

### Body surface potentials recording and ECG electrodes localization

A routine clinical resting 12-lead electrocardiogram (ECG) was recorded during the study visit, and ECG metrics were measured by the 12SL algorithm (GE Marquette Electronics, Milwaukee, WI).

Unipolar ECG potentials were recorded on the body surface using the ActiveTwo biopotential measurement system (BioSemi, Amsterdam, the Netherlands) with 128 Ag/AgCL electrodes (4 panels of 32 electrodes; each panel is arranged as four strips of 8-electrodes; diameter of the ECG electrodes 5 mm).^11^ The sampling rate of the signal was 16,384 Hz; bandwidth DC-3,200 Hz. ECG electrodes were localized by three-dimensional (3D) photography approach, using a PeacsKinect (Peacs BV, Arnhem, the Netherlands) and Kinect camera (Microsoft, Redmond, WA, USA).^11^ For co-registration of torso images, five CMR- or CT-specific markers were placed on each patient’s chest to wear during scanning, to mark ECG electrode locations.

### Reconstruction of torso and heart meshes

We constructed 3D meshes of a continuous surface of the endocardium (excluding papillary muscles) and epicardium of both ventricular chambers. The 3D heart and torso meshes were reconstructed using a semi-automatic approach—image growing method of a continuous surface^11, 12^ from CMR/CT images using ITK-snap software (PICSL, USA).^13^ Each cardiac mesh was manually reviewed to ensure a continuous segmentation of epicardium and endocardium of both ventricular chambers and exclude atria chambers and papillary muscles. Both torso meshes segmented by the 3D photography method and DICOM images were matched using the co-registered CMR/CT markers and electrode position, as previously described.^11^ The resolution of the cardiac mesh was 3.6 ± 0.5 mm with 3992 ± 735 nodes.

### Inverse solution and reconstruction of the cardiac activation map

The workflow is shown in Figure 1. One clean normal sinus beat was selected for analysis; an absence of extrasystole before and after the selected beat was verified. We used the open-source SCIRun problem-solving environment developed at the Center for Integrative Biomedical Computing (University of Utah, UT),^14, 15^ which was previously used to compute forward and inverse solutions^16^ and reconstruct unipolar epicardial and endocardial electrograms (EGMs).

**Figure 1.**
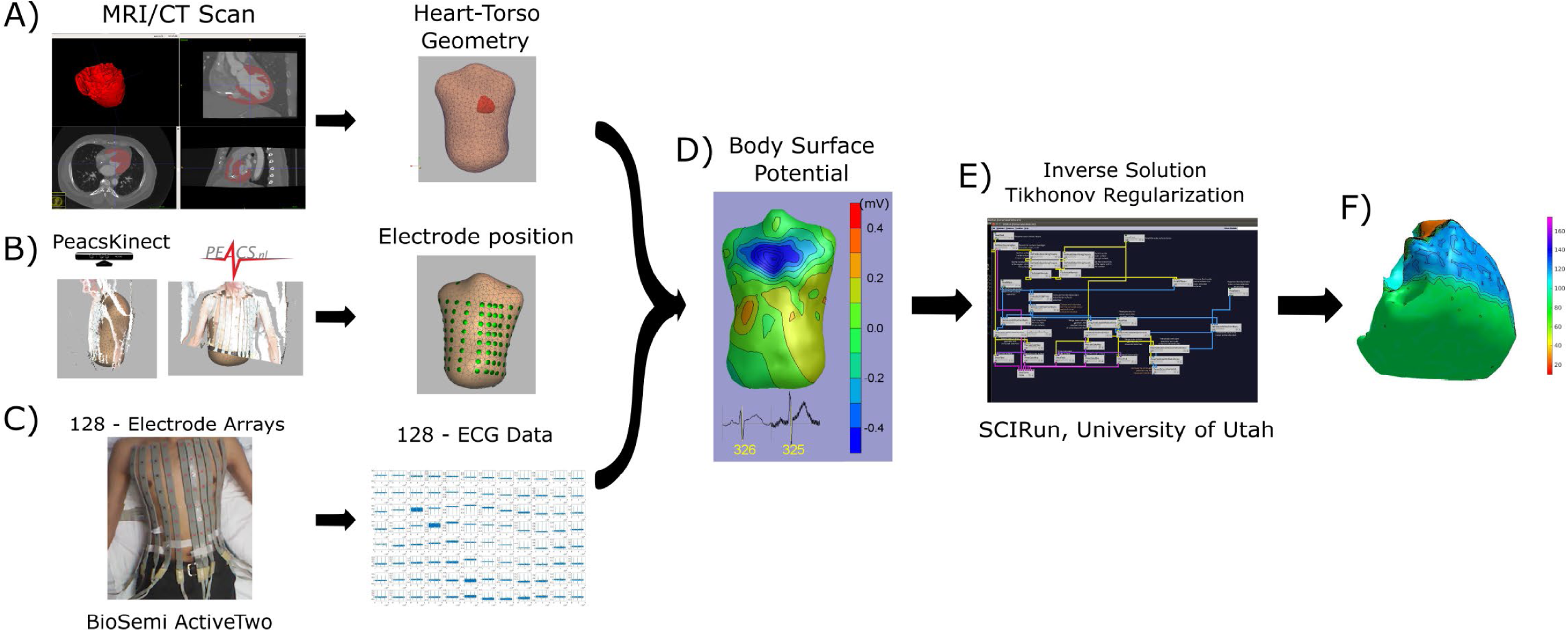
Workflow from data acquisition to data analysis. (A) MRI/CT scan to obtain heart and torso 3D meshes; (B) electrode position from kinect camera using PeacsKinect, and (C) 128-ECG recordings using Biopac system, were used to obtain a (D) body surface potential, and (E) reconstruction of ventricular potential map using SCI-run toolkit..

The inverse problem was solved as the potential-based formulation (boundary element method), as a weighted minimum norm problem by applying a Tikhonov L2-norm regularization. The source code and inverse problem SCIRun toolkit with documentation can be found at www.scirun.org.

The steepest downslope of each unipolar EGM was determined automatically, using MATLAB (The MathWorks Inc, Natick, MA) software application. In order to verify the consistency of morphology and the steepest downslope detection, each pair of neighboring unipolar EGMs—together with the resulting bipolar EGM (calculated as their difference)—were reviewed by at least two investigators (AW, KY, NMR, EAPA) who were blinded to the study group assignment(Figure 2). In the case of disagreement between all 3 EGMs, the unipolar EGMs were excluded from further analysis. Recalculation of the steepest downslope was performed in the case of morphology agreement but steepest downslope disagreement. For calculation of the time reference point, the three limb leads (I, II, and III) were used to define the average QRS onset on the surface ECG. Local activation time (LAT) in each node of the mesh was calculated as the time difference between average (surface ECG) QRS onset and the time of the steepest downslope (minimum dV/dt) on a corresponding unipolar EGM. To reconstruct the cardiac activation map on the epicardial and endocardial surface, the LAT was plotted in each epicardial and endocardial node.

**Figure 2.**
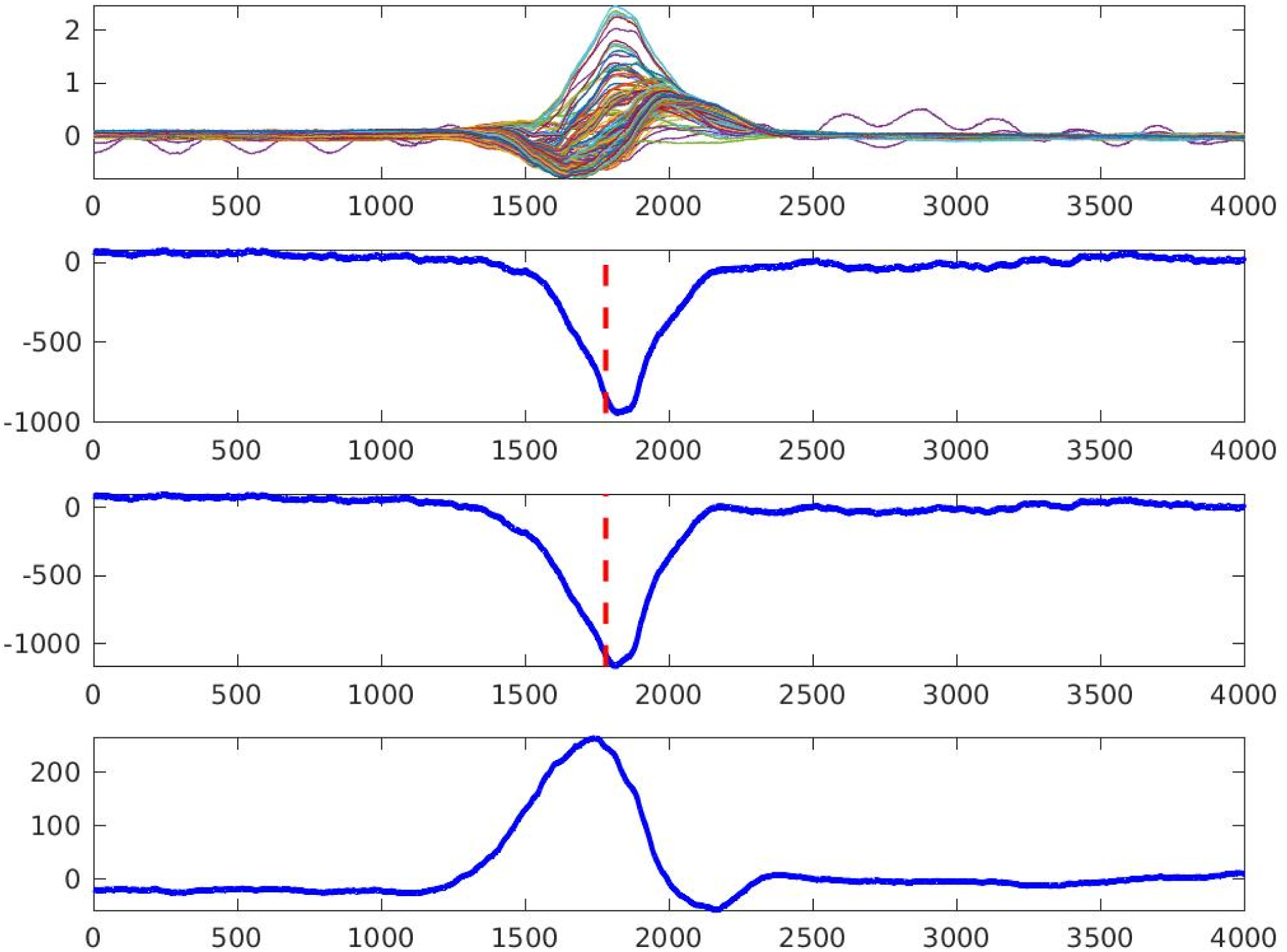
Quality control of the measurement of the steepest downslope of epicardial electrogram (the steepest negative dV/dt, dashed line). Two neighboring unipolar electrograms (two middle rows) and bipolar electrogram (bottom row) are shown. An upper row shows overlapped 128 body surface electrocardiograms. Local activation time is marked by the dashed line.

### Unipolar voltage potential map

Unipolar voltage was measured in each reconstructed EGM, and the unipolar voltage potential maps were constructed. The peak-to-peak voltage on each unipolar EGM was automatically measured using a MATLAB (The MathWorks Inc, Natick, MA) software application. Investigators (AW, KY, NMR, EAPA), blinded to the group assignment, validated the accuracy of the detection of unipolar EGM peaks. We used the standardized myocardial segmentation and nomenclature^9^ to define 17 segments of the LV. The mean unipolar voltage was calculated for each segment. RV endocardial surface in 5 segments (basal anteroseptal and inferoseptal, mid-cavity anteroseptal and inferoseptal, and apical septal) served as an “epicardial” surface of the LV. Standard deviation (SD) of unipolar voltage distribution in each segment served as a measure of voltage dispersion within each segment.

### Dispersion of local activation time

Mean LAT was calculated for each LV segment.^9^ RV endocardial surface in 5 segments (basal anteroseptal and inferoseptal, mid-cavity anteroseptal and inferoseptal, and apical septal) served as an “epicardial” surface of the LV. The dispersion of activation was measured as SD of LAT in each segment.

### Statistical analysis

Statistics of normally distributed variables are summarized as mean±SD. Distributions of all variables were reviewed. After verifying the normality of distribution, we tested the hypothesis that the mean voltage is the same across four study groups while removing the assumption of equal covariance matrices. The Wald chi-squared statistic with James’s approximation^17^ was used to calculate P-values.

We used a Kruskal–Wallis test of the hypothesis that four study groups are from the same population, to compare voltage and LAT dispersions (measured as an SD of LAT and voltage in each of 17 segments^9^), which have a non-normal distribution. Non-normally distributed variables are summarized as the median and interquartile range (IQR). A P-value of less than 0.05 was considered significant. Statistical analyses were performed using STATA MP 15.1 (StataCorp LLC, College Station, TX).

## Results

### Study population

The clinical characteristics of the study population are shown in Table 1. Most of HCM patients (80%) had previously undergone genetic testing. The definitive, disease-causing *MYHBPC3* mutation was found in 2 patients. Half of the HCM participants had survived a sudden cardiac arrest, and the other half had a documented history of sustained VT. While half of the HCM patients had a history of severe LVOT obstruction (up to 153 mmHg at peak exertion), they had already undergone surgical myectomy, resulting in vastly improved LVOT gradients (provoked peak LVOT gradient 20±20 mmHg), at the time of enrollment.

**Table 1.** Demographic and clinical characteristics of the study population

| **Characteristic** | **Healthy (n=10)** | **Post-MI VT-free (n=10)** | **Post-MI VT (n=10)** | **HCM (n=10)** |
| --- | --- | --- | --- | --- |
| Age(SD), y | 28.8(5.6) | 69.3(9.3) | 64.0(10.5) | 61.3(9.3) |
| Male, n(%) | 4(40) | 10(100) | 10(100) | 8(80) |
| White, n(%) | 6(60) | 10(100) | 9(90) | 10(100) |
| BMI(SD), kg/m<sup>2</sup> | 23.6(3.6) | 33.0(6.6) | 30.9(6.9) | 30.1(5.7) |
| History of hypertension, n(%) | 0 | 9(90) | 9(90) | 6(60) |
| History of diabetes, n(%) | 0 | 5(50) | 2(20) | 1(10) |
| Chronic kidney disease, n(%) | 0 | 3(30) | 0 | 0 |
| Class I antiarrhythmics, n(%) | 0 | 0 | 5(50) | 2(20) |
| Beta-blockers, n(%) | 0 | 7(70) | 9(90) | 8(80) |
| ACEI/ARB/ARNI | 0 | 6(60) | 8(80) | 2(20) |
| Aldosterone antagonists, n(%) | 0 | 4(40) | 2(20) | 0 |
| Diuretics, n(%) | 0 | 8(80) | 2(20) | 0 |
| LVEF(SD), % | 65.8(4.7) | 45.8(15.4) | 37.3(12.7) | 60.4(8.3) |
| GLS(SD), % | - | -13.5(0) | -9.6(0.4) | -13.2(0.8) |
| LVIDd(SD), cm | 4.3(0.4) | 5.2(1.3) | 6.1(1.1) | 4.5(0.7) |
| LVIDs(SD), cm | 2.7(0.4) | 4.2(0.9) | 5.1(1.4) | 2.9(0.4) |
| LAVI(SD), ml/m<sup>2</sup> | 22.3(3.1) | 32.6(13.9) | 33.2(8.8) | 40.1(12.8) |
| LVEDVI(SD), ml/m<sup>2</sup> | 76.7(13.4) | 89.6(30.,0) | 100.6(34.1) | 72.2(12.3) |
| RVEDVI(SD), ml/m<sup>2</sup> | 89.0(20.5) | 71.8(14.4) | 76.7(10.8) | 76.3(13.4) |
| LV mass index(SD),g/m<sup>2</sup> | 63.9(13.9) | 76.0(15.0) | 92.2(25.7) | 99.3(28.9) |
| **LVH, %** | 0 | 1 | 6 | 6 |
| IVSd(SD), cm | 0.8(0.1) | 1.5(1.4) | 1.0(0.1) | 1.5(0.3) |
| LVPWd(SD), cm | 0.8(0.1) | 1.1(0.2) | 1.0(0.2) | 1.2(0.3) |
| E/A ratio | 1.7(0.2) | 1.3(0.8) | 0.9(0.2) | 1.0(0.4) |
| E/e' ratio | 5.9(1.5) | 9.1(3.4) | 11.4(5.7) | 9.5(4.0) |
| Peak LVOT gradient(SD), mmHg | 4.7(1.1) | 6.2(2.2) | 10.2(9.0) | 77.9(193) |
| LVOT diameter(SD), cm | 2.2(0.1) | 2.2(0.2) | 2.3(0.3) | 2.2(0.3) |
| Wall motion score(SD) | 8.8(8.9) | 27.4(16.6) | 18.2(14.4) | 10.3(8.9) |
| Resting heart rate(SD), bpm | 76.8(13.7) | 74.2(11.6) | 66.9(15.1) | 70.7(10.5) |
| QTc interval(SD), ms | 408(24) | 398(32) | 444(23) | 424(37) |

LVEF=left ventricular ejection fraction; GLS=global longitudinal strain; LVIDd = Left ventricular internal dimension at end-diastole; LVIDs = Left ventricular internal dimension at end-systole; LVEDV = left ventricular end-diastolic volume; LVEDVI = left ventricular end-diastolic volume index; RVEDV = right ventricular end-diastolic volume; RVEDVI = right ventricular end-diastolic volume index; IVSd=Interventricular septum thickness at end-diastole; LVPWd=Left ventricular posterior wall thickness at end-diastole; LAVI = left atrial volume index; SD = standard deviation; LVH=left ventricular hypertrophy

Healthy controls had no comorbidities and did not take medications (Table 1). Nearly all group II-III participants and HCM patients had hypertension, but only a minority of participants had diabetes and chronic kidney disease. Nearly all post-MI and HCM patients were on beta-blockers. Half of group III participants and 20% of HCM group were taking antiarrhythmic medications (amiodarone, sotalol, mexiletine).

In the VT-free post-MI group, the scar was located in the anteroseptal region in 90% of participants. In post-MI VT group, the scar was located in the inferoposterior region in 40% and anteroseptal in 60%. A single-chamber ICD was implanted in approximately 50% of patients. The other half had a dual-chamber ICD implanted.

LV systolic function was normal in healthy controls and HCM participants, whereas ischemic cardiomyopathy (ICM) with reduced LVEF was confirmed for both post-MI groups. LVH was equally present in HCM and ICM with VT groups.

### Mean unipolar voltage and unipolar voltage dispersion

A representative example of voltage maps is shown in Figure 3. Mean unipolar voltage (Table 2 and Figure 4) was significantly different across all four study groups in basal anteroseptal and apical septal segments on both sides of septum – LV and RV endocardium. Also, a significant difference in voltage across all four groups was observed on both endocardial and epicardial surfaces of basal anterior and anterolateral segments, the endocardial surface of anterior apical segment, and the epicardial surface of basal inferior and mid-inferolateral segments. Healthy individuals had the smallest mean unipolar voltage, whereas HCM was characterized by the largest voltage (Figure 4). Unipolar voltage in the two post-MI groups was similar, and had intermediate values, as compared to healthy and HCM participants.

**Table 2.** Mean (SD) unipolar voltage potentials (µV) in LV endocardial and epicardial, and RV endocardial septal regions.

| Region |  | Healthy (n=10) | Post-MI VT-free (n=10) | Post-MI VT (n=10) | HCM (n=10) | P<sub>James</sub> |
| --- | --- | --- | --- | --- | --- | --- |
| Endocardial | Basal anterior | **802(108)** | **853(281)** | **982(230)** | **1086(241)** | **0.016** |
|  | Basal anteroseptal | **812(128)** | **931(304)** | **954(166)** | **1085(266)** | **0.043** |
|  | Basal inferoseptal | 829(103) | 901(359) | 943(318) | 1121(239) | **0.071** |
|  | Basal inferior | 823(106) | 804(275) | 942(241) | 1037(276) | 0.130 |
|  | Basal inferolateral | 856(98) | 835(291) | 958(209) | 1081(286) | 0.125 |
|  | Basal anterolateral | **829(82)** | **949(337)** | **967(259)** | **1096(300)** | **0.023** |
|  | Mid-anterior | 838(114) | 871(323) | 945(187) | 1045(214) | **0.090** |
|  | Mid-anteroseptal | 817(98) | 903(291) | 900(211) | 1042(276) | 0.136 |
|  | Mid-inferoseptal | 839(107) | 901(332) | 916(224) | 1087(290) | 0.135 |
|  | Mid-inferior | 793(83) | 852(281) | 952(262) | 1008(251) | **0.071** |
|  | Mid-inferolateral | 836(98) | 890(305) | 926(260) | 1111(297) | **0.086** |
|  | Mid-anterolateral | 837(94) | 936(414) | 984(238) | 1110(244) | 0.204 |
|  | Apical anterior | **809(111)** | **984(452)** | **969(154)** | **1094(314)** | **0.036** |
|  | Apical septal | **810(133)** | **898(450)** | **941(205)** | **1122(229)** | **0.028** |
|  | Apical Inferior | 835(97) | 863(282) | 902(233) | 1015(211) | 0.198 |
|  | Apical lateral | 791(143) | 892(295) | 964(181) | 942(180) | 0.133 |
| Epicardial / RV endocardial | Basal anterior | **842(90)** | **898(358)** | **934(189)** | **1094(211)** | **0.023** |
|  | RV<sub>endo</sub> Basal anteroseptal | **827(157)** | **929(326)** | **961(174)** | **1120(217)** | **0.028** |
|  | RV<sub>endo</sub> Basal inferoseptal | 808(133) | 883(325) | 904(234) | 1073(302) | 0.251 |
|  | Basal inferior | **838(99)** | **904(280)** | **964(228)** | **1053(177)** | **0.028** |
|  | Basal inferolateral | 847(113) | 903(322) | 936(177) | 1069(205) | **0.064** |
|  | Basal anterolateral | **855(131)** | **937(355)** | **996(166)** | **1125(218)** | **0.027** |
|  | Mid-anterior | 856(118) | 901(307) | 942(182) | 1106(242) | **0.068** |
|  | RV<sub>endo</sub> Mid-anteroseptal | 824(118) | 912(415) | 911(185) | 1068(288) | 0.201 |
|  | RV<sub>endo</sub> Mid-inferoseptal | 812(136) | 893(349) | 895(154) | 975(231) | 0.320 |
|  | Mid-inferior | 838(88) | 920(335) | 966(229) | 1002(211) | 0.142 |
|  | Mid-inferolateral | **829(114)** | **904(341)** | **968(201)** | **1098(197)** | **0.013** |
|  | Mid-anterolateral | 856(107) | 875(296) | 941(165) | 1076(215) | **0.069** |
|  | Apical anterior | **825(102)** | **883(288)** | **950(195)** | **1075(241)** | **0.042** |
|  | RV<sub>endo</sub> Apical septal | **833(97)** | **937(365)** | **902(129)** | **1078(166)** | **0.008** |
|  | Apical Inferior | 829(102) | 838(263) | 933(166) | 1024(188) | **0.068** |
|  | Apical lateral | 825(100) | 884(360) | 918(206) | 1082(266) | **0.070** |
|  | Apex | 840(94) | 888(320) | 935(182) | 1054(217) | **0.069** |

**Figure 3.**
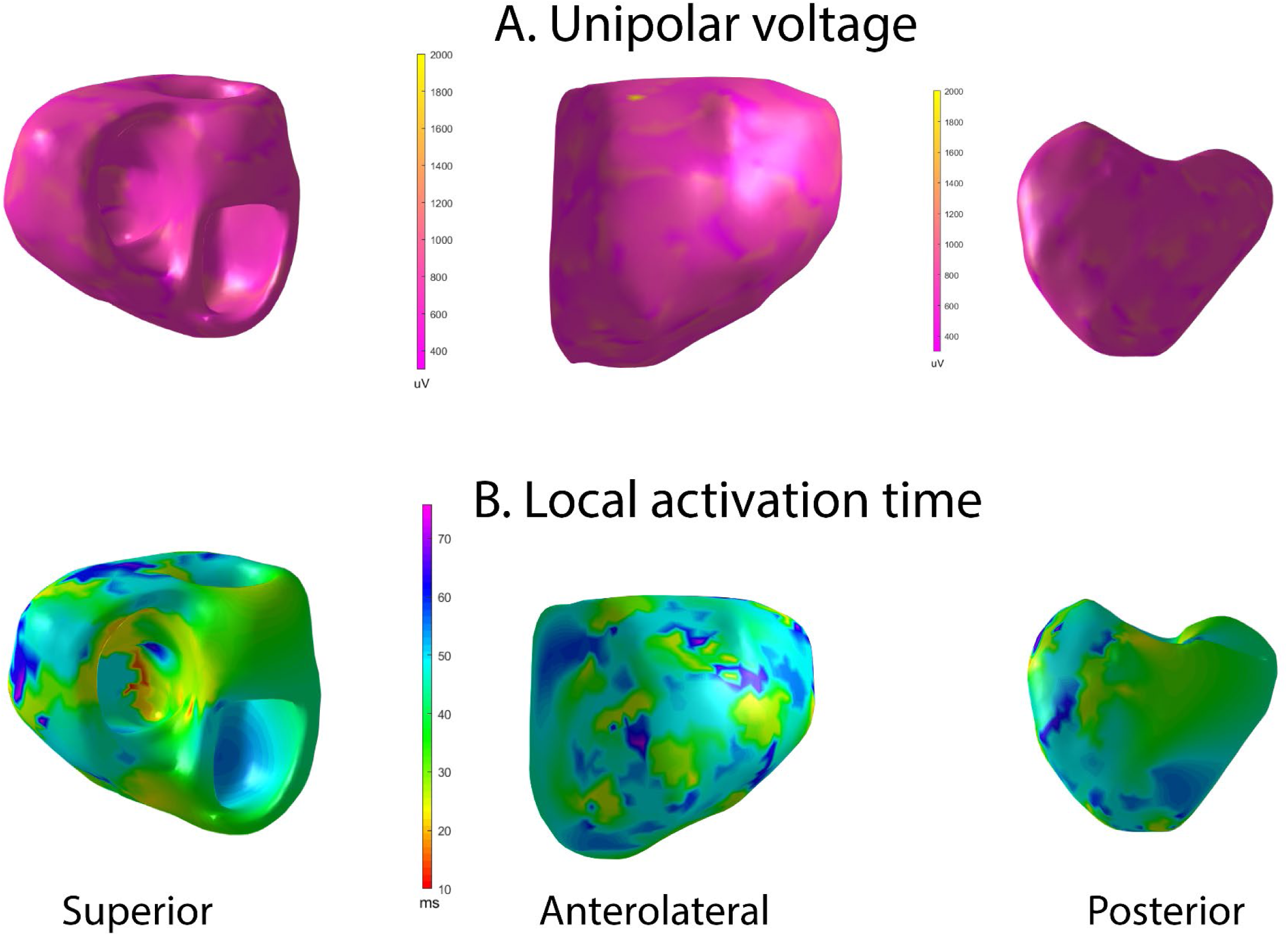
Representative examples of (A) unipolar voltage and (B) ventricular activation maps in a healthy participant, during sinus rhythm activation. Superior, anterolateral, and a posterior view. The orifices of the aorta and the mitral valve are combined. Supplemental movies 1 and 2 show representative examples of unipolar voltage and ventricular activation maps in a healthy participant, during sinus rhythm activation.

**Figure 4.**
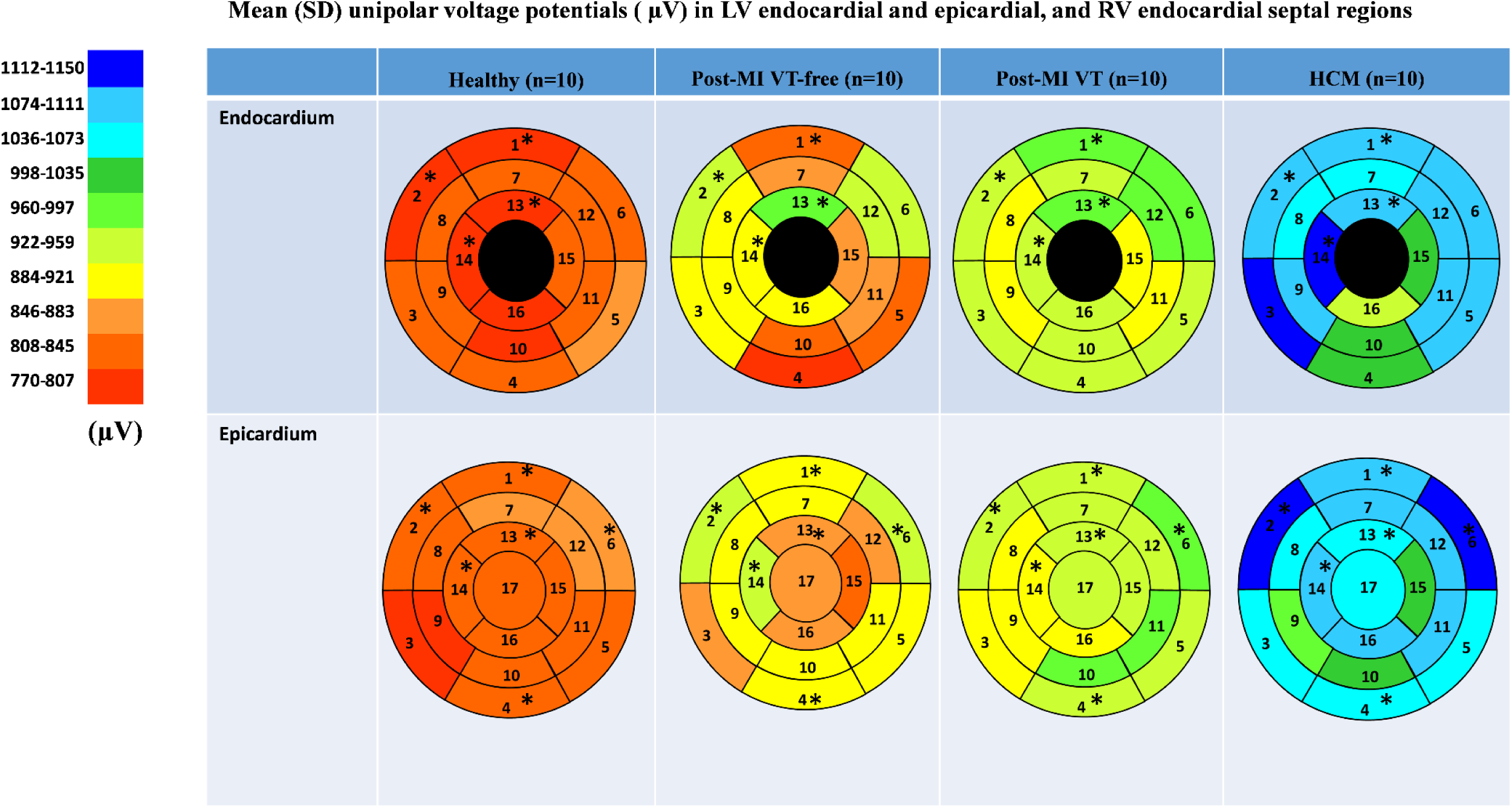
Comparison of mean unipolar voltage in study groups, in 16 LV endocardial and 17 LV epicardial segments. Nomenclature: 1=basal anterior; 2=basal anteroseptal; 3=Basal inferoseptal; 4=Basal inferior; 5=Basal inferolateral; 6=Basal anterolateral; 7=Mid-anterior; 8=Mid-anteroseptal; 9=Mid-inferoseptal; 10=Mid-inferior; 11=Mid-inferolateral; 12=Mid-anterolateral; 13=Apical anterior; 14=Apical septal; 15=Apical Inferior; 16=Apical lateral; 17=apex. RV endocardial surface in 5 segments (basal anteroseptal and inferoseptal, mid-cavity anteroseptal and inferoseptal, and apical septal) served as an “epicardial” surface of the LV. Statistically significant differences are marked with a star.

Voltage dispersion was significantly smaller in healthy, as compared to the other three groups (Table 3 and Figure 5). Remarkably, in several segments, voltage dispersion in HCM was the highest amongst all four groups, significantly exceeding voltage dispersion in both ICM groups. The unipolar voltage dispersion was significantly different across study groups in both endocardial and epicardial segments of the basal anterior, basal anterolateral and inferolateral, apical inferior, and the epicardial surface of the apex.

**Table 3.** Unipolar voltage dispersion within segments on endocardial and epicardial surface of the left ventricle

|  | Region | Healthy (n=10) | Post-MI VT-free (n=10) | Post-MI VT (n=10) | HCM (n=10) | P<sub>Kruskal-Wallis</sub> |
| --- | --- | --- | --- | --- | --- | --- |
| Endocardial | Basal anterior | 101(53-147) | 161(102-214) | 146(89-157) | 178(162-276) | **0.017** |
|  | Basal anteroseptal | 73(42-132) | 94(78-266) | 148(64-227) | 223(166-295) | 0.053 |
|  | Basal inferoseptal | 104(75-129) | 123(84-199) | 130(59-152) | 230(155-314) | 0.066 |
|  | Basal inferior | 95(54-126) | 126(79-152) | 147(111-203) | 184(130-267) | 0.076 |
|  | Basal inferolateral | 88(80-117) | 114(89-153) | 105(87-170) | 177(124-252) | **0.031** |
|  | Basal anterolateral | 58(49-117) | 153(128-195) | 142(94-215) | 201(136-224) | **0.049** |
|  | Mid-anterior | 82(73-105) | 167(100-241) | 127(80-229) | 169(138-224) | 0.113 |
|  | Mid-anteroseptal | 50(36-59) | 80(50-136) | 127(48-171) | 112(47-290) | 0.345 |
|  | Mid-inferoseptal | 87(54-117) | 84(36-148) | 125(66-204) | 187(106-227) | 0.277 |
|  | Mid-inferior | 113(69) | 111(79-285) | 169(122-180) | 183(72-196) | 0.752 |
|  | Mid-inferolateral | 110(48-176) | 139(120-282) | 158(112-197) | 109(24-188) | 0.368 |
|  | Mid-anterolateral | 105(92-121) | 112(83-196) | 136(73-190) | 219(48-335) | 0.588 |
|  | Apical anterior | 67(18-125) | 141(76-170) | 184(87-224) | 248(102-306) | 0.160 |
|  | Apical septal | 107(106-127) | 120(83-154) | 170(113-233) | 180(110-260) | 0.222 |
|  | Apical Inferior | 63(11-109) | 151(97-229) | 63(61-141) | 155(106-199) | **0.035** |
|  | Apical lateral | 66(34-94) | 160(102-202) | 159(121-221) | 63(23-163) | 0.097 |
| Epicardial / RV endocardial | Basal anterior | 110(106-168) | 158(109-236) | 189(143-208) | 215(161-281) | **0.041** |
|  | RV Basal anteroseptal | 112(105-150) | 162(99-233) | 152(91-201) | 259(176-311) | 0.109 |
|  | RV Basal inferoseptal | 105(71-122) | 135(90-229) | 202(106-243) | 172(117-319) | 0.116 |
|  | Basal inferior | 106(90-131) | 179(146-323) | 202(112-223) | 207(130-311) | **0.019** |
|  | Basal inferolateral | 107(98-121) | 151(131-185) | 198(118-210) | 185(167-290) | **0.004** |
|  | Basal anterolateral | 120(101-147) | 145(135-161) | 166(153-200) | 172(140-268) | **0.025** |
|  | Mid-anterior | 150(128-163) | 170(128-235) | 195(166-234) | 201(168-338) | 0.060 |
|  | RV Mid-anteroseptal | 106(49-134) | 180(119-225) | 150(112-191) | 165(102-280) | 0.187 |
|  | RV Mid-inferoseptal | 90(69-123) | 153(89-182) | 136(92-167) | 89(59-208) | 0.338 |
|  | Mid-inferior | 132(88-155) | 178(116-260) | 182(173-197) | 217(151-277) | **0.040** |
|  | Mid-inferolateral | 79(75-134) | 129(118-255) | 190(132-228) | 180(150-320) | **0.024** |
|  | Mid-anterolateral | 138(125-153) | 145(99-144) | 167(117-203) | 217(156-248) | 0.260 |
|  | Apical anterior | 158(109-182) | 163(124-275) | 180(119-226) | 220(175-244) | 0.185 |
|  | RV Apical septal | 129(95-149) | 182(128-261) | 194(161-219) | 175(124-272) | 0.070 |
|  | Apical Inferior | 127(106-143) | 150(128-179) | 135(105-198) | 222(152-261) | **0.047** |
|  | Apical lateral | 115(107-129) | 126(105-207) | 146(106-164) | 182(156-341) | 0.229 |
|  | Apex | 118(100-129) | 166(123-274) | 206(130-229) | 260(163-315) | **0.019** |

**Figure 5.**
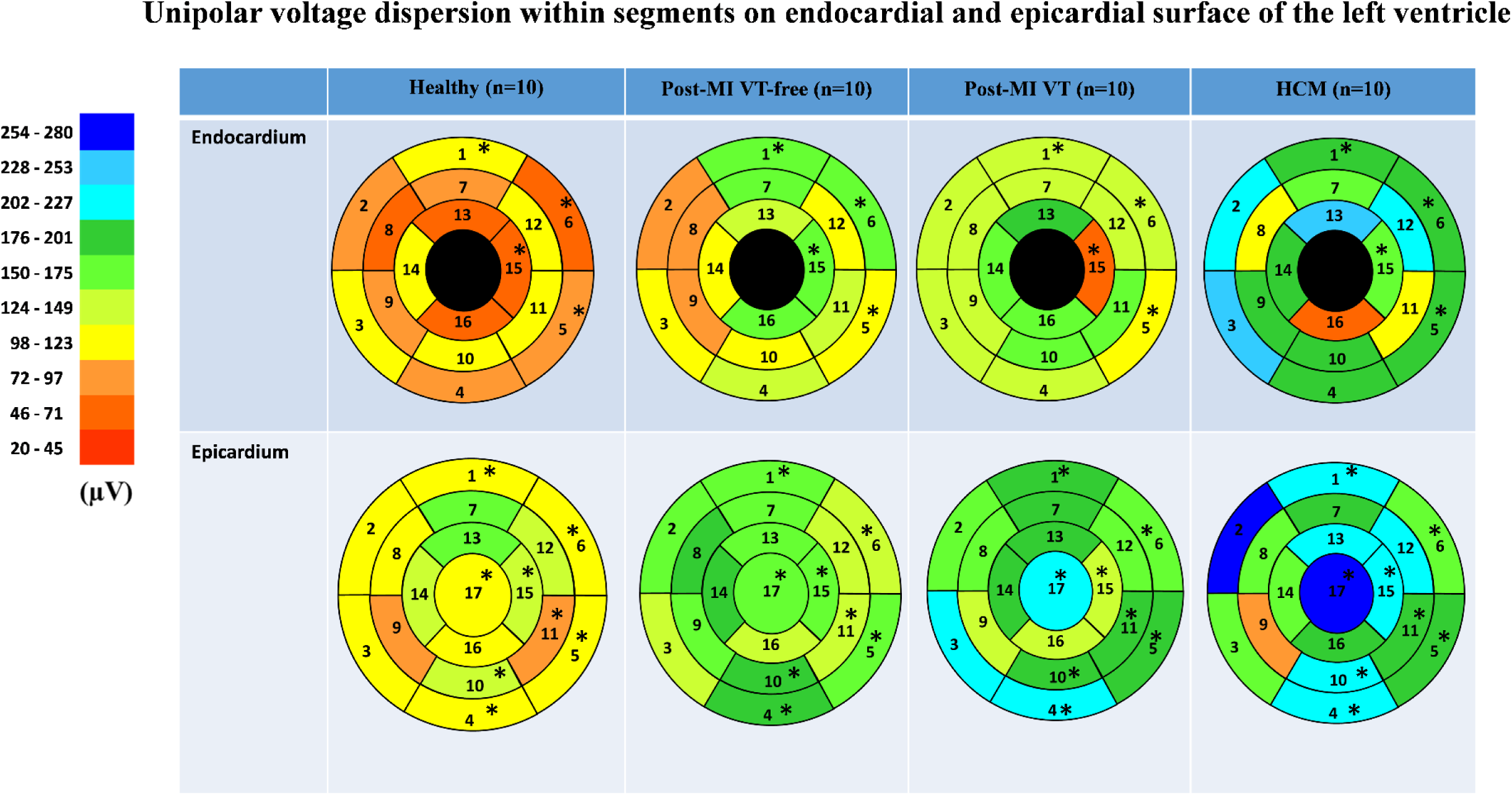
Comparison of unipolar voltage dispersion in study groups, in 16 LV endocardial and 17 LV epicardial segments. Segment nomenclature is described in the Figure 4 legend. Statistically significant differences are marked with a star.

### Dispersion of ventricular activation

A representative example of the activation map is shown in Figure 3. In Healthy, we observed a normal activation pattern, which initiated in the septal region and propagated from endocardium to epicardium, with several breakthroughs – near the RV apex and anterior paraseptal aspects of the epicardium in regions adjacent to the left anterior descendent coronary artery. Activation proceeded from apex to the inferior basal area in both RV and LV, with the inferolateral LV base and the region near the right ventricular outflow tract (RVOT) being the latest to activate.

The dispersion of LAT was significantly higher in HCM patients in nearly all epicardial basal segments and basal lateral endocardial segments (Table 4 and Figure 6). A consistent trend in LAT dispersion between groups was observed, with the smallest LAT dispersion in healthy subjects, intermediate LAT dispersion in ICM patients, and the largest LAT dispersion in HCM patients. Furthermore, in ICM and HCM patients there was a trend towards greater LAT dispersion in basal segments, as compared to apical or mid-segments, whereas in healthy controls, small degree of LAT dispersion was homogeneous, without apex-base gradient. In addition, there was a trend of larger LAT dispersion in post-MI patients with VT history, as compared to VT-free post-MI patients.

**Table 4.** Local activation dispersion in msec (median, interquartile range) on left ventricular endocardial and epicardial surface

|  | Region | Healthy (n=10) | Post-MI VT-free (n=10) | Post-MI VT (n=10) | HCM (n=10) | P<sub>Kruskal-Wallis</sub> |
| --- | --- | --- | --- | --- | --- | --- |
| Endocardial surface | Basal anterior | 8.9(6.9-10.9) | 9.8(5.9-13.5) | 9.5(5.7-16.0) | 16.0(10.7-17.2) | 0.084 |
|  | Basal anteroseptal | 5.8(5.2-12.0) | 10.6(8.7-13.3) | 12.6(11.2-15.7) | 13.3(5.2-21.1) | 0.473 |
|  | Basal inferoseptal | 7.7(5.2-9.5) | 11.8(9.7-17.9) | 12.5(7.7-16.2) | 15.8(10.0-18.8) | 0.053 |
|  | Basal inferior | 6.4(4.2-9.1) | 9.8(5.9-13.4) | 12.7(3.8-16.0) | 14.6(11.5-15.3) | 0.082 |
|  | Basal inferolateral | 6.9(6.3-11.8) | 7.0(3.2-16.0) | 12.0(7.2-14.9) | 16.5(14.3-20.1) | **0.014** |
|  | Basal anterolateral | 7.0(6.4-9.6) | 10.2(9.0-12.3) | 10.5(7.5-16.1) | 17.3(12.1-22.2) | **0.009** |
|  | Mid-anterior | 8.1(6.1-9.2) | 11.0(5.5-16.8) | 10.2(6.1-14.9) | 11.1(8.6-13.0) | 0.402 |
|  | Mid-anteroseptal | 7.5(2.7-8.8) | 8.4(5.4-11.8) | 11.6(6.6-14.6) | 10.1(5.7-14.7) | 0.248 |
|  | Mid-inferoseptal | 8.9(5.8-13.4) | 6.5(4.8-12.3) | 9.3(4.0-10.8) | 11.0(6.9-11.8) | 0.854 |
|  | Mid-inferior | 11.1(2.9-13.4) | 8.9(5.8-12.9) | 8.6(3.4-12.2) | 9.2(3.8-12.3) | 0.924 |
|  | Mid-inferolateral | 5.5(4.5-9.3) | 11.2(8.6-14.1) | 6.3(3.9-8.9) | 8.5(1.9-13.4) | 0.252 |
|  | Mid-anterolateral | 6.6(5.3-9.2) | 7.7(5.5-9.3) | 9.7(3.8-16.1) | 9.6(4.8-14.0) | 0.676 |
|  | Apical anterior | 4.2(0.7-14.8) | 9.0(5.6-11.2) | 9.3(7.6-13.4) | 9.3(6.5-10.1) | 0.507 |
|  | Apical septal | 5.3(2.7-17.6) | 7.8(5.8-14.1) | 7.8(6.2-11.3) | 10.5(9.1-14.1) | 0.727 |
|  | Apical Inferior | 6.2(2.1-8.4) | 11.1(9.6-14.1) | 6.9(2.9-14.2) | 7.4(1.2-10.1) | 0.062 |
|  | Apical lateral | 7.0(1.4-13.6) | 8.8(1.9-12.7) | 10.1(3.4-18.2) | 4.5(1.3-9.9) | 0.454 |
| Epicardial | Basal anterior | 8.3(7.7-9.8) | 12.8(11.4-15.9) | 13.2(11.9-15.1) | 13.5(12.4-22.3) | **0.0007** |
|  | RV Basal anteroseptal | 8.1(7.0-11.3) | 11.1(9.3-12.8) | 14.1(8.9-15.4) | 17.7(14.4-22.0) | **0.012** |
|  | RV Basal inferoseptal | 9.8(7.4-12.9) | 15.7(8.3-16.5) | 10.3(9.6-15.7) | 12.7(9.7-18.3) | 0.676 |
|  | Basal inferior | 8.5(7.7-9.9) | 12.6(11.7-17.5) | 12.5(11.5-14.1) | 16.3(11.4-17.5) | **0.0008** |
|  | Basal inferolateral | 11.1(7.7-11.9) | 12.6(10.8-13.9) | 14.2(12.2-16.6) | 17.1(14.7-20.2) | **0.006** |
|  | Basal anterolateral | 9.9(9.2-11.7) | 10.7(9.1-14.9) | 13.8(11.1-16.0) | 17.5(16.0-24.4) | **0.002** |
|  | Mid-anterior | 8.9(8.3-11.0) | 10.7(7.9-15.3) | 10.2(8.0-12.0) | 12.4(10.1-14.2) | 0.364 |
|  | RV Mid-anteroseptal | 8.9(6.3-10.4) | 9.1(7.1-12.5) | 11.8(7.3-14.6) | 11.6(6.9-15.5) | 0.720 |
|  | RV Mid-inferoseptal | 9.4(3.3-11.9) | 6.4(4.7-10.9) | 9.4(5.9-12.0) | 2.7(0.5-8.2) | 0.676 |
|  | Mid-inferior | 9.8(9.3-11.1) | 9.3(7.8-11.5) | 9.8(7.9-14.8) | 9.9(6.9-13.6) | 0.977 |
|  | Mid-inferolateral | 10.5(7.7-11.8) | 9.1(7.3-13.8) | 11.4(7.1-12.2) | 13.6(8.3-16.3) | 0.558 |
|  | Mid-anterolateral | 9.3(7.3-10.6) | 9.5(6.6-12.1) | 8.4(7.0-13.9) | 11.3(10.3-12.8) | 0.429 |
|  | Apical anterior | 8.9(8.0-10.9) | 7.9(7.5-9.4) | 11.4(7.9-12.4) | 9.6(8.8-13.6) | 0.257 |
|  | RV Apical septal | 6.4(5.3-8.5) | 7.6(6.0-10.0) | 9.4(6.8-13.8) | 11.6(8.9-12.2) | 0.060 |
|  | Apical Inferior | 9.9(8.1-10.9) | 8.5(6.4-10.7) | 10.1(8.5-12.9) | 10.0(9.0-14.3) | 0.443 |
|  | Apical lateral | 8.3(5.4-9.6) | 8.2(6.1-9.6) | 9.0(7.9-10.9) | 9.4(7.1-12.1) | 0.730 |
|  | Apex | 8.0(7.4-10.3) | 7.3(7.0-8.7) | 9.2(7.1-10.0) | 11.0(9.9-12.8) | 0.051 |

**Figure 6.**
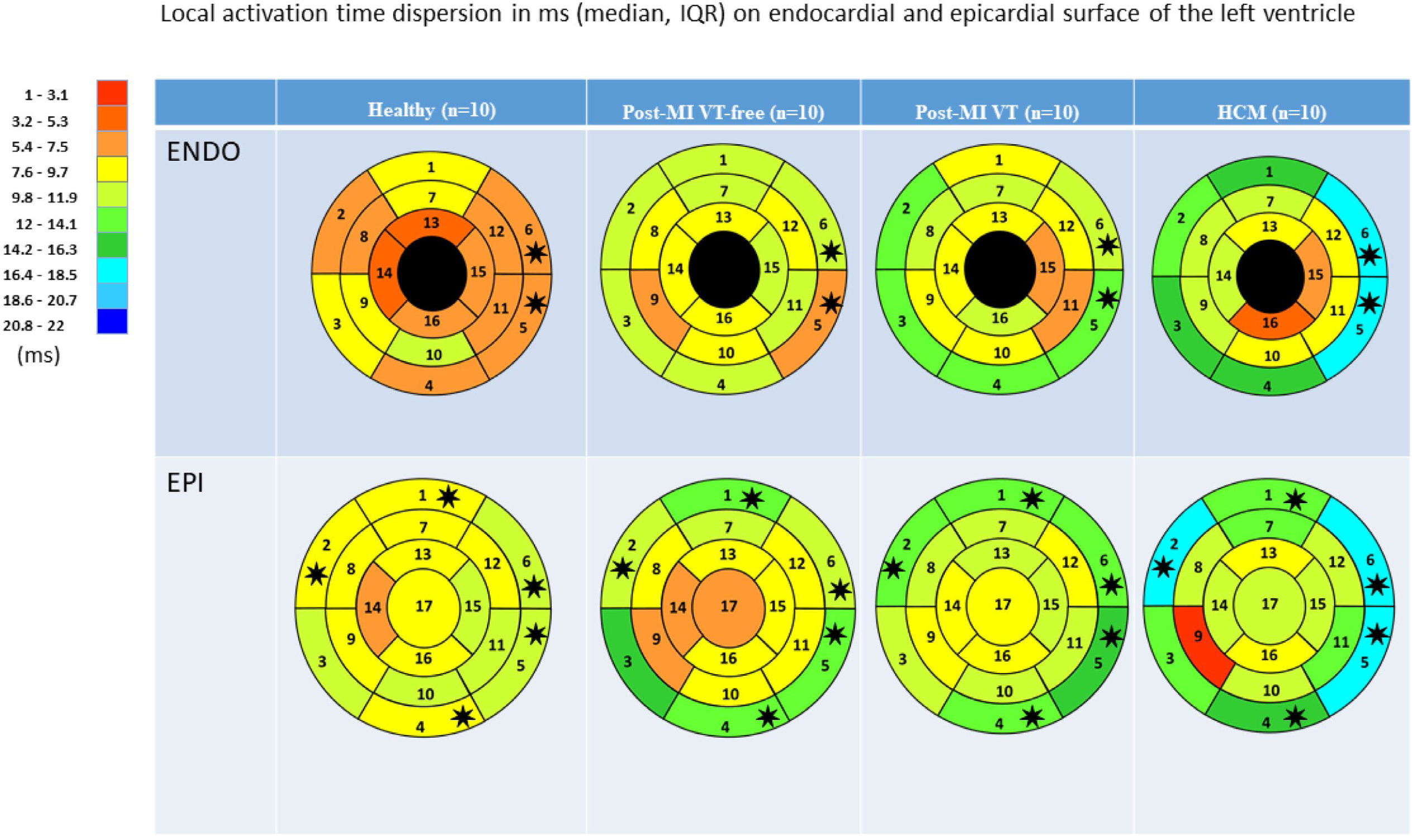
Comparison of ventricular activation dispersion in study participants, in 16 LV endocardial and 17 LV epicardial segments. Segment nomenclature is described in the Figure 4 legend. Statistically significant differences are marked with a star.

## Discussion

Our study revealed the features of the EP substrate in HCM, which differentiate the HCM substrate from the ICM substrate in patients with VT history and post-MI scar located in the same areas (anterior and anteroseptal segments). HCM is characterized by a greater degree of activation dispersion in basal segments, an apex-base gradient in activation dispersion, a larger voltage and a greater voltage dispersion. Dispersion of ventricular activation contributes to the dispersion of total recovery time and facilitates the development and maintenance of reentrant VT.^18^ Dispersion of ventricular activation is a well-known mechanism of reentrant VT ^18^, which can explain substantial risk of VT and sudden cardiac death in HCM

### EP substrate and mechanisms of arrhythmogenesis in HCM

HCM is the most common monogenic cardiac disease.^19^ Diagnosis of HCM is challenging, although some preliminary machine learning studies are promising.^20, 21^ Prior cellular studies demonstrated that enhanced late sodium current is an important mechanism of arrhythmogenesis in HCM ^2^. Enhanced late sodium current in HCM cardiomyocytes manifested by action potential prolongation and increased frequency of early depolarizations and delayed afterdepolarizations, suggesting triggered mechanism of VT, similar to that in long QT syndromes. However, the RESTYLE-HCM randomized controlled trial ^22^ did not demonstrate a benefit of the late sodium channel blocker ranolazine in symptomatic patients with nonobstructive HCM.

The results of our study suggest explanation to ineffectiveness of pharmacological late sodium current blockade in HCM. In 1988, Vassallo et al^18^ showed differences between post-infarction scar-related reentrant VT and triggered VT in long QT syndrome. Patients with post-infarction scar and reentrant VT mechanism manifested dispersion of endocardial activation, rather than dispersion of refractoriness. In contrast, patients with long QT syndrome and triggered VT mechanism manifested dispersion of refractoriness, rather than dispersion of activation. The results of our study demonstrated dispersion of activation in HCM patients, supporting the presence of electrophysiological substrate of reentrant VT, resembling VT mechanism in post-infarction patients. HCM is characterized by a disorganized sarcomeric alignment, which can augment nonuniform anisotropic conduction, creating a substrate for reentry (both slow conduction and unidirectional block).^23^ Disorganized bundles of ventricular fibers can lead to asymmetry in conduction. An impulse conducting in one direction meets a different sequence of muscle branching and changes in muscle bundle diameter, as compared to an impulse conducting in the opposite direction. Such asymmetry affects the source-sink relationships.^24^

We showed a greater degree of voltage dispersion in HCM as compared to post-MI patients in both endocardial and the epicardial segments in basal anterior, basal anterolateral and inferolateral, apical inferior, and the epicardial surface of the apex. This finding may be explained by an underlying phenomenon of diffused interstitial fibrosis in HCM, generating greater voltage dispersion as compared to patchy post-MI fibrosis. Further MRI studies^25^ utilizing late gadolinium enhancement and T1-mapping are needed to evaluate the agreement between voltage dispersion on ECGi voltage maps and imaging-defined type of fibrosis, and their associations with clinical outcomes in HCM patients.

Very few previous studies reported results of electroanatomical mapping in HCM. Reported findings included local conduction delay or conduction block, fractionated electrograms, and reduced voltage.^26^ Scattered intramural fibrosis in HCM did not manifest by the low voltage on endocardial, nor epicardial unipolar maps. However, Schumacher et al^26^ reported reduced bipolar voltage in the septal region in HCM patients. In this study, we observed significantly larger unipolar voltage in HCM as compared to healthy persons or ICM patients, and the difference was especially prominent in HCM-affected regions of the heart: both endocardial and epicardial basal and apical septal segments. The different methodology of voltage mapping (unipolar vs. bipolar) may explain the observed differences between our results and previous contact mapping studies. A previous HCM case report^27^ describing ECGi findings did not provide results of the voltage map from their HCM patient. Consistent with our findings, Yoshida et al^28^ in 1986 conducted body surface isopotential mapping and showed that HCM patients have significantly larger peak-to-peak voltage than patients with LVH due to essential hypertension.

### Noninvasive mapping of ventricular activation

In this study, we used the Forward/Inverse problem toolkit from the SCIRun problem-solving environment, which is used by many investigators in the field.^14, 15, 29, 30^ However, knowing the limitations of the ECGi method,^31^ we intentionally limited our analysis by averaged “per segment” data. Duchateau et al^31^ observed a mean activation time error of approximately 20 ms. Nevertheless, in this study, we observed statistically significant differences in ventricular activation dispersion. In the base of LV, dispersion of activation was approximately twice larger in HCM patients, as compared to healthy controls. In spite of limitations, our findings of a higher voltage dispersion through entire LV and an apex-base gradient in increased activation dispersion in HCM provided meaningful insight into mechanisms of arrhythmogenesis in HCM. Further development of ECGi method is needed.

### Limitations

A case-control study is susceptible to bias. We selected only high-risk HCM patients, and our HCM case sample may not be representative of all HCM patients. To minimize selection bias, all groups were enrolled in the same single center. Further development of inverse solution noninvasive activation mapping method is needed to enable interpretation of local ventricular activation patterns. The study size was small. Validation of the study findings in larger studies is needed. Because of the high-risk study population, we did not design the discontinuation of medications during the study. It is known that the use of antiarrhythmic medications may affect ventricular conduction and results of the body surface mapping. Nevertheless, our study allowed group comparison of contemporary patient population on guideline-recommended medical therapy. Genetic testing was not performed for all study participants. As the prevalence of HCM gene carriers in the general population was estimated at 0.5% ^19^, a low chance exists that group I-III participants carry HCM gene. However, only 10% of group II participants, and no control participants had LVH, which supports the study validity.

## Data Availability

n/a

## Acknowledgments

The authors thank the study participants and staff. We thank William Woodward, ARMRIT, for the help with the CMR data acquisition. The study was funded by Gilead Sciences, Inc, as a physician-initiated study (LGT). This work was partially supported by 1R01HL118277 and 2R56HL118277 (LGT).

## Authors contribution statement

EP run inverse solutions computations and built activation and voltage maps. KH computed mean (SD) LAT and voltage per segment, and prepared figures. DG, CF, and MF helped to design the study and analyzed imaging data. CH, JT, EP, and LT collected the study data. KJ, FP, NR, KY, and AW analyzed the ECG and EGM data. KD and SH helped to design the study and facilitated enrollment. All authors drafted the study and revised the manuscript. LT designed the study, handled the study funding, directed the study implementation, including quality assurance and control, conducted statistical analyses, and critically revised the manuscript. All authors approved the submitted version and have agreed both to be personally accountable for the author’s own contributions and the accuracy and integrity of any part of the work.

